# Comparative Clinical Outcomes of Everolimus versus Biolimus-Eluting Stents: A Meta-Analysis of 27,071 Patients from Randomized Trials

**DOI:** 10.1101/2025.09.29.25336296

**Authors:** I M Khalid Reza, Shankar Biswas, Elangovan Krishnan, KC Anil, Varshitha Bezawada, Spurthy Sri Namala, Mansi Chaudhari, Joel Jayan Peniel, Zainab Abid Shah, Yashvant Pravinbhai Bodar, Hakim Ullah Wazir, Somaiya Ahmed, Okasha Tahir, Jeimy Marilyn Castellanos, Mohammad Semaal Khan

## Abstract

**Background:** The comparative clinical outcomes of everolimus-eluting stents (EES) versus biolimus-eluting stents (BES) remains uncertain despite widespread use. This study conducted a systematic review and meta-analysis of randomized controlled trials comparing contemporary drug-eluting stent platforms.

**Methods:** We searched MEDLINE, Embase, CENTRAL, and Scopus through May 2025 for randomized trials comparing EES with BES in patients undergoing percutaneous coronary intervention. The primary outcomes were major adverse cardiac events (MACE) and device-oriented composite endpoint (DOCE) at the longest follow-up. Secondary outcomes included all-cause mortality, target lesion revascularization (TLR), and stent thrombosis. Random-effects meta-analysis was performed using risk ratios (RR) and 95% confidence intervals (CI). Heterogeneity was assessed using I² statistics. The GRADE approach evaluated the certainty of evidence.

**Results:** Thirteen trials randomizing 27,071 patients (12,226 EES; 14,845 BES), with follow-up 9-120 months, were included. EES demonstrated a trend toward reduced MACE compared with BES (9.7% vs 10.3%; RR 0.93, 95% CI: 0.87-1.00; p=0.053; I²=0%) with an absolute risk reduction of 0.7% (number needed to treat=142). DOCE showed similar results (RR 0.94, 95% CI: 0.88-1.01; p=0.10; I²=0%). No significant differences were observed for all-cause mortality (RR 0.96, 95% CI: 0.88-1.05; p=0.37), TLR (RR 0.95, 95% CI: 0.86-1.05; p=0.36), or stent thrombosis (RR 0.93, 95% CI: 0.69-1.23; p=0.60), all with I²=0%. Subgroup analysis by follow-up duration showed consistent results (p-interaction=0.88). No publication bias was detected. GRADE assessment indicated moderate certainty evidence for most outcomes.

**Conclusions:** In this meta-analysis, EES showed a trend toward reduced MACE compared with BES, though not reaching statistical significance. The remarkable homogeneity and small absolute differences suggest both platforms provide excellent outcomes. Both stent platforms remain appropriate first-line options for percutaneous coronary intervention.

**Registration:** PROSPERO CRD42025108092

## INTRODUCTION

Percutaneous coronary intervention with drug-eluting stents (DES) has become the dominant revascularization strategy for patients with coronary artery disease, with over 3 million procedures performed annually worldwide [1]. The evolution from bare-metal stents to first-generation DES dramatically reduced restenosis rates but was associated with concerns about late stent thrombosis, prompting the development of second-generation devices with improved safety profiles [2]. Contemporary DES platforms have largely overcome these early limitations, achieving target lesion revascularization rates below 5% and stent thrombosis rates under 1% at 1 year [3].

Among second-generation DES, everolimus-eluting stents (EES) and biolimus-eluting stents (BES) represent two distinct design philosophies that have gained widespread clinical adoption. EES platforms, including the Xience (Abbott Vascular) and Promus (Boston Scientific) families, utilize everolimus—a sirolimus analog with potent antiproliferative properties—delivered from a durable fluorinated copolymer applied to cobalt-chromium or platinum-chromium struts of 74-81 μm thickness [4]. The durable polymer provides controlled drug release with 80% elution within 30 days, while maintaining long-term vessel wall coverage [4].

In contrast, BES platforms such as Nobori (Terumo) and BioMatrix (Biosensors) incorporate biolimus A9, a highly lipophilic sirolimus analog specifically developed for stent applications [5]. A distinguishing characteristic of BES is the biodegradable poly-lactic acid polymer applied only to the abluminal surface, which completely degrades within 6-9 months, theoretically reducing long-term inflammatory responses [4]. However, BES platforms utilize thicker struts (112-120 μm) on stainless steel backbones, which may influence procedural and clinical outcomes [6].

The comparative clinical outcomes of these two design approaches remains uncertain. Previous meta-analyses have compared biodegradable polymer DES as a class against durable polymer DES, yielding conflicting results. Bangalore et al. reported increased mortality with biodegradable polymer DES beyond 1 year, while El-Hayek et al. found no significant differences in clinical outcomes [7,8]. However, these analyses combined heterogeneous DES platforms with different drugs, polymers, and strut designs, potentially masking true differences between specific stent types. The network meta-analysis by Palmerini et al. suggested superiority of cobalt-chromium EES over BES [9], but indirect comparisons are subject to transitivity assumptions that may not hold across diverse trial populations.

Direct randomized comparisons between EES and BES have been conducted across various clinical settings and geographic regions, but individual trials have been underpowered to detect differences in clinical endpoints. The NEXT trial demonstrated non-inferiority of BES compared with EES at 1 year in 3,235 Japanese patients [10], while the COMPARE II trial showed similar results in 2,707 all-comer patients [11]. Long-term follow-up from these trials has become available [10,12], along with additional randomized comparisons, providing an opportunity for a comprehensive synthesis of the evidence.

Furthermore, the optimal duration of dual antiplatelet therapy remains a topic of debate for biodegradable versus durable polymer DES [13]. Although shorter durations may be feasible with biodegradable polymers due to improved healing, current guidelines do not differentiate recommendations based on polymer type [14]. Understanding the comparative safety of EES versus BES, particularly regarding stent thrombosis risk, is essential for individualizing antiplatelet therapy duration.

Therefore, we conducted a systematic review and meta-analysis of randomized controlled trials directly comparing everolimus-eluting stents with biolimus-eluting stents in patients undergoing percutaneous coronary intervention. Our objective was to evaluate the long-term efficacy and safety of EES and BES stent platforms in contemporary clinical practice and to provide evidence-based guidance for stent selection.

## METHODS

### Protocol and Registration

This systematic review and meta-analysis was conducted according to a pre-specified protocol developed in accordance with the Preferred Reporting Items for Systematic Review and Meta-Analysis Protocols (PRISMA-P) guidelines [15] and reported in accordance with the PRISMA 2020 statement [16]. The protocol was registered with PROSPERO (CRD42025108092).

### Eligibility Criteria

#### Study Design

We included randomized controlled trials comparing everolimus-eluting stents (EES) with biolimus-eluting stents (BES) in patients undergoing percutaneous coronary intervention, with a minimum follow-up of 30 days. Both superiority and non-inferiority trial designs were eligible.

#### Participants

Adult patients (≥18 years) undergoing percutaneous coronary intervention were included regardless of clinical presentation (stable coronary artery disease or acute coronary syndromes). Studies involving native coronary arteries and saphenous vein grafts were eligible.

#### Interventions

The intervention group included any everolimus-eluting stent platform (Xience, Promus, Synergy). The comparator group included any biolimus-eluting stent platform (Nobori, BioMatrix). Only direct head-to-head comparisons were included.

#### Outcomes

Primary outcomes were: (1) major adverse cardiac events (MACE), variably defined across studies but generally including death, myocardial infarction, and revascularization; and (2) device-oriented composite endpoint (DOCE), defined as cardiac death, target vessel myocardial infarction, and target lesion revascularization. Secondary outcomes included all-cause mortality, target lesion revascularization, and definite or probable stent thrombosis per Academic Research Consortium definitions [17].

### Information Sources and Search Strategy

We searched PubMed (n = 84), Embase (n = 587), CENTRAL (n = 163), and Scopus (n = 421) from their inception through May 2025. The search combined terms for everolimus, biolimus, drug-eluting stents, and randomized trials without language restrictions. The detailed search strategies for each database are provided in Supplementary Table S1.

### Study Selection

Two reviewers independently screened 720 unique records after removing 535 duplicates from 1,255 total identified records. Full-text assessment was performed for 21 articles. Disagreements were resolved through consensus. We excluded 8 articles: no direct EES vs BES comparison (n=2), subgroup comparison (n=1), and conference paper (n=5). Details of the excluded studies and reasons for exclusion are provided in Supplementary Table S2.

### Data Extraction

Two reviewers independently extracted data using standardized forms. Extracted by data included study characteristics, patient demographics, intervention details, and outcome events. For the 13 included trials, data were available for 27,071 patients (12,226 EES; 14,845 BES) for MACE analysis. When trials reported multiple follow-up periods, we used the longest available (range: 9-120 months). We attempted to extract bleeding outcomes and detailed DAPT protocols but these were inconsistently reported across trials.

### Risk of Bias Assessment

Risk of bias was assessed using the Cochrane RoB 2 tool [18,19] across five domains. Ten studies were judged to have a low overall risk of bias, two studies had some concerns, and one study had a high risk of bias due to substantial loss to follow-up. All studies employed blinded endpoint adjudication committees.

### Data Synthesis and Statistical Analysis

#### Effect Measures

Risk ratios with 95% confidence intervals were calculated for all dichotomous outcomes using intention-to-treat data.

### Synthesis Methods

Random-effects meta-analysis was performed using the DerSimonian-Laird method [20]. Analyses were conducted using R version 4.3.2 [21] with the meta package. The actual analysis included:

- 14 comparisons from 13 trials for MACE (27,071 patients)
- 10 comparisons for DOCE (24,939 patients)
- 13 comparisons for mortality (26,870 patients)
- 11 comparisons for TLR (25,140 patients)
- 12 comparisons for stent thrombosis (26,710 patients

### Heterogeneity Assessment

Heterogeneity was assessed using I² statistics and Q-test [22]. The observed I² was 0% for all primary analyses, indicating no statistical heterogeneity.

### Additional Analyses

### Subgroup Analyses

We performed subgroup analysis by follow-up duration (≤1 year: 5 studies; 1-5 years: 6 studies; >5 years: 3 studies) with formal tests for interaction.

### Sensitivity Analyses

We conducted: (1) leave-one-out analysis excluding each study sequentially [23]; (2) cumulative meta-analysis by publication year [24]; (3) trim-and-fill analysis for publication bias [25].

### Meta-regression

Random-effects meta-regression examined the influence of follow-up duration, diabetes prevalence (19-46% range), ACS percentage (16-100% range), and publication year (2011-2023) on treatment effects.

### Assessment of Publication Bias

Publication bias was assessed through funnel plot inspection and Egger’s test for outcomes with ≥10 studies. Trim-and-fill analysis was performed to adjust for potential missing studies.

### Assessment of Reporting Bias

Reporting bias was assessed by examining trial registrations (when available) and comparing prespecified outcomes with those reported in published manuscripts. We evaluated whether all outcomes mentioned in the methods sections were adequately reported in the results and whether any additional outcomes not prespecified were included.

### Quality of Evidence Assessment

Grading of Recommendations Assessment, Development and Evaluation (GRADE) methodology [19] was applied to assess the certainty of evidence for each outcome. Starting from high certainty for randomized controlled trials, we evaluated five domains for potential downgrading: risk of bias, inconsistency, indirectness, imprecision, and publication bias. Each domain could lead to downgrading by one or two levels, resulting in final ratings of high, moderate, low, or very low certainty. Final assessments were: moderate certainty for MACE, mortality, and TLR (downgraded for risk of bias); low certainty for stent thrombosis (downgraded for risk of bias and imprecision).

## RESULTS

### Study Selection and Characteristics

The systematic literature search identified 1,255 records through electronic databases **(Figure 1)**. After removing 535 duplicates, 720 records underwent title and abstract screening, of which 699 were excluded. Full-text assessment of 21 articles resulted in the exclusion of 8 studies for the following reasons: no direct EES versus BES comparison (n=2), subgroup comparison (n=1), and conference paper (n=5). Ultimately, 13 randomized controlled trials met the inclusion criteria **(Table 1)**, comprising 27,071 patients (12,226 EES; 14,845 BES) for the primary outcome analysis.

**Figure 1.**
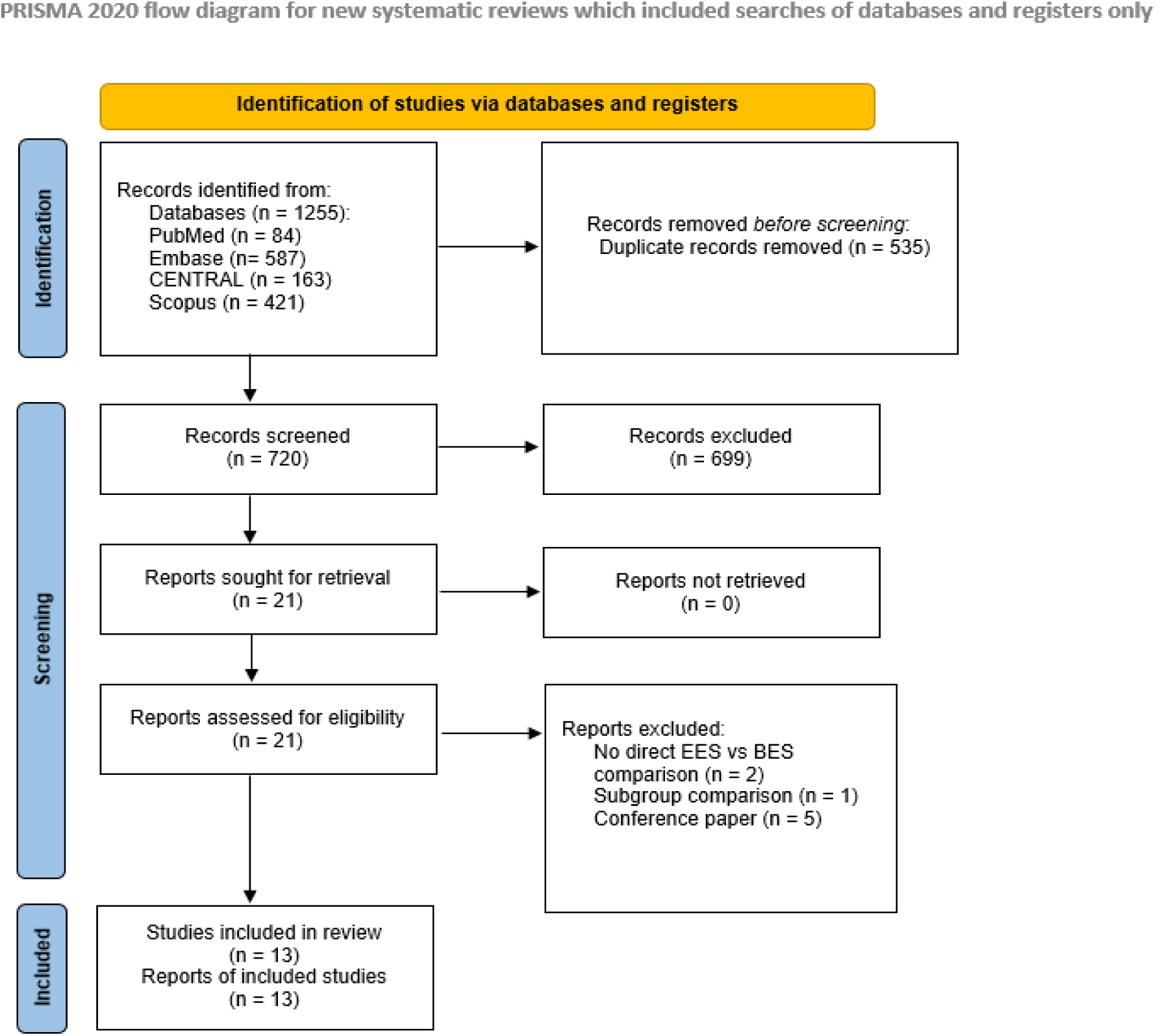
PRISMA Flow Diagram. Study selection process according to PRISMA (Preferred Reporting Items for Systematic Reviews and Meta-Analyses) 2020 guidelines. The systematic search identified 1,255 records, of which 13 randomized controlled trials comparing everolimus-eluting stents with biolimus-eluting stents met the inclusion criteria for the meta-analysis.

**Table 1.** Characteristics of Included Randomized Controlled Trials.

| Study<br>(Reference) | Follow-up<br>period<br>(months) | Trial name | Country | Sample<br>size |  | Age (years) |  | Male (%) |  | Primary<br>Endpoint | Industry<br>Funded | Risk<br>of<br>Bias |
| --- | --- | --- | --- | --- | --- | --- | --- | --- | --- | --- | --- | --- |
|  |  |  |  | EE<br>S | BE<br>S | EES | BES | EES | BE<br>S |  |  |  |
| Jakl<br>2023<br>[26] | 9<br>60* | ROBUST | Czech<br>Republic<br>, United<br>States | 100 | 101 | 59.2±1<br>0 | 58.4±<br>9.6 | 82 | 89 | OCT<br>strut<br>coverage | Yes | Low |
| Maeng<br>2019<br>[32] | 12 | SORT<br>OUT VIII | Denmark | 138<br>5 | 137<br>9 | 66±11 | 66±1<br>1 | 77 | 77 | TLF | Yes | Low |
| Lee 2014<br>[28] | 12 | LONG-DES V | South<br>Korea | 255 | 245 | 63.5±1<br>0.6 | 63.1±<br>10.5 | 72.2 | 68.2 | In-<br>segment<br>LLL | Yes | High |
| Natsuaki<br>2013<br>[10] | 12 | NEXT | Japan | 161<br>8 | 161<br>7 | 69.3±9<br>.8 | 69.1±<br>9.8 | 77 | 77 | TLR | Yes | Low |
| Natsuaki<br>2015<br>[27] | 36 | NEXT | Japan | 161<br>8 | 161<br>7 | 69.3±9<br>.8 | 69.1±<br>9.8 | 77 | 77 | TLR | Yes | Low |
| Natsuaki<br>2023<br>[31] | 120 | NEXT | Japan | 121<br>3 | 120<br>4 | 69.5±9<br>.7 | 69±9.<br>8 | 77 | 77 | TLF | Yes | Some<br>concerns |
| Puricel<br>2015<br>[33] | 9 | EVERBIO<br>II | Switzerla<br>nd | 80 | 80 | 65±11 | 65±1<br>0 | 80 | 80 | LLL | Yes | Low |
| Kaiser<br>2015<br>[34] | 24 | BASKET-PROVE II | Europe | 765 | 765 | 62±11 | 62±1<br>1 | 80 | 78 | Cardiac<br>death/MI | No | Low |
| Youn<br>2020<br>[35] | 24 | CHOICE | South<br>Korea | 638 | 634 | 65.6±1<br>1 | 65±1<br>1.6 | 66.1 | 70.2 | TLF | No | Some<br>concerns |
| Smits<br>2013<br>[11] | 12 | COMPAR<br>E II | Europe | 912 | 179<br>5 | 62.7±1<br>1 | 63±1<br>1.1 | 74.3 | 74.4 | MACE | Yes | Low |
| Vlachojannis<br>2017<br>[12] | 60 | COMPAR<br>E II | Europe | 912 | 179<br>5 | 62.7±1<br>1 | 63±1<br>1.1 | 74.3 | 74.4 | MACE | Yes | Low |
| Vlachojannis<br>2017<br>[30] | 36 | NEXT +<br>COMPAR<br>E II | Japan,<br>Europe | 253<br>0 | 341<br>2 | 66.9±1<br>0.8 | 65.9±<br>10.9 | 76.3 | 75.6 | Composite | Yes | Low |
| Separham<br>2011<br>[36] | 12 | - | Iran | 100 | 100 | 62.4±1<br>0.2 | 60.6±<br>9.1 | 64 | 66 | MACE | No | Low |

\* Long-term follow-up
Abbreviations: EES, everolimus-eluting stent; BES, biolimus-eluting stent; OCT, optical coherence tomography; TLR, target lesion revascularization; LLL, late lumen loss; TLF, target lesion failure; MI, myocardial infarction; MACE, major adverse cardiac events.

The included trials were published between 2011 and 2023, with sample sizes ranging from 201 to 3,412 patients per trial **(Table 1)**. Follow-up duration varied from 9 to 120 months, with 5 trials reporting outcomes of at least 3 years [26,27]. Ten trials (83%) were industry-funded. All trials employed 1:1 randomization except COMPARE II (2:1; BES:EES), and 11 (92%) were designed as non-inferiority studies. The mean follow-up completeness was 94.3% (range: 92.5-98.5%).

### Patient and Procedural Characteristics

The mean age of enrolled patients ranged from 58.8 to 69.3 years (weighted mean 64.7 years), with male predominance ranging from 65% to 86%. The prevalence of diabetes mellitus varied from 19% to 46%, while the proportion of patients presenting with acute coronary syndromes ranged from 16% to 100%, with one trial exclusively enrolling STEMI patients [26].

Regarding stent platforms, the EES group predominantly utilized Xience/Promus family stents (Abbott Vascular/Boston Scientific) with strut thickness ranging from 74 to 81 μm. The BES group primarily employed Nobori (Terumo; n=8 trials) or BioMatrix (Biosensors; n=5 trials) platforms with a strut thickness of 112-120 μm. Notably, 12 of 13 trials (92%) used durable polymer EES, while all BES platforms utilized biodegradable polymers. The mean number of stents implanted per patient was 1.5±0.6, with a mean total stent length of 32.4±18.2 mm. Dual antiplatelet therapy duration ranged from 6 to 12 months across trials. The technical specifications and procedural characteristics of the stent platforms are summarized in **Table 3**.

### Risk of Bias Assessment

Risk of bias assessment using the RoB 2 tool revealed generally high-quality evidence (Supplementary Figure 1). Among the 13 included RCTs, 10 studies (76.9%) demonstrated low overall risk of bias, 2 studies (15.4%) had some concerns, and 1 study (7.7%) showed high risk of bias. All studies exhibited low risk for randomization, outcome measurement, and selective reporting domains. The primary methodological concern was deviations from intended interventions (9/13 studies with some concerns due to the open-label design inherent to device trials). One study had high risk due to missing outcome data (substantial loss of angiographic follow-up) [28]. The predominantly low risk of bias across studies supports the reliability of our findings.

### Primary Outcomes

#### Major Adverse Cardiac Events (MACE)

Fourteen comparisons from 13 trials comprising 27,071 patients reported MACE outcomes. Contributing studies included a mix of all-comer populations and specific subgroups, with follow-up ranging from 9 to 120 months. Most studies (10/13, 76.9%) had low risk of bias, with consistent endpoint definitions across trials. The incidence of MACE was 9.7% (1,192/12,226) in the EES group versus 10.3% (1,534/14,845) in the BES group. Meta-analysis demonstrated a risk ratio (RR) of 0.93 (95% confidence interval [CI]: 0.87-1.00; p=0.053) favoring EES, with no evidence of heterogeneity (I²=0%, p=0.62) **(Figure 2)**. The absolute risk reduction was 0.7%, corresponding to a number needed to treat (NNT) of 142 (95% CI: 73 to 12,677).

**Figure 2.**
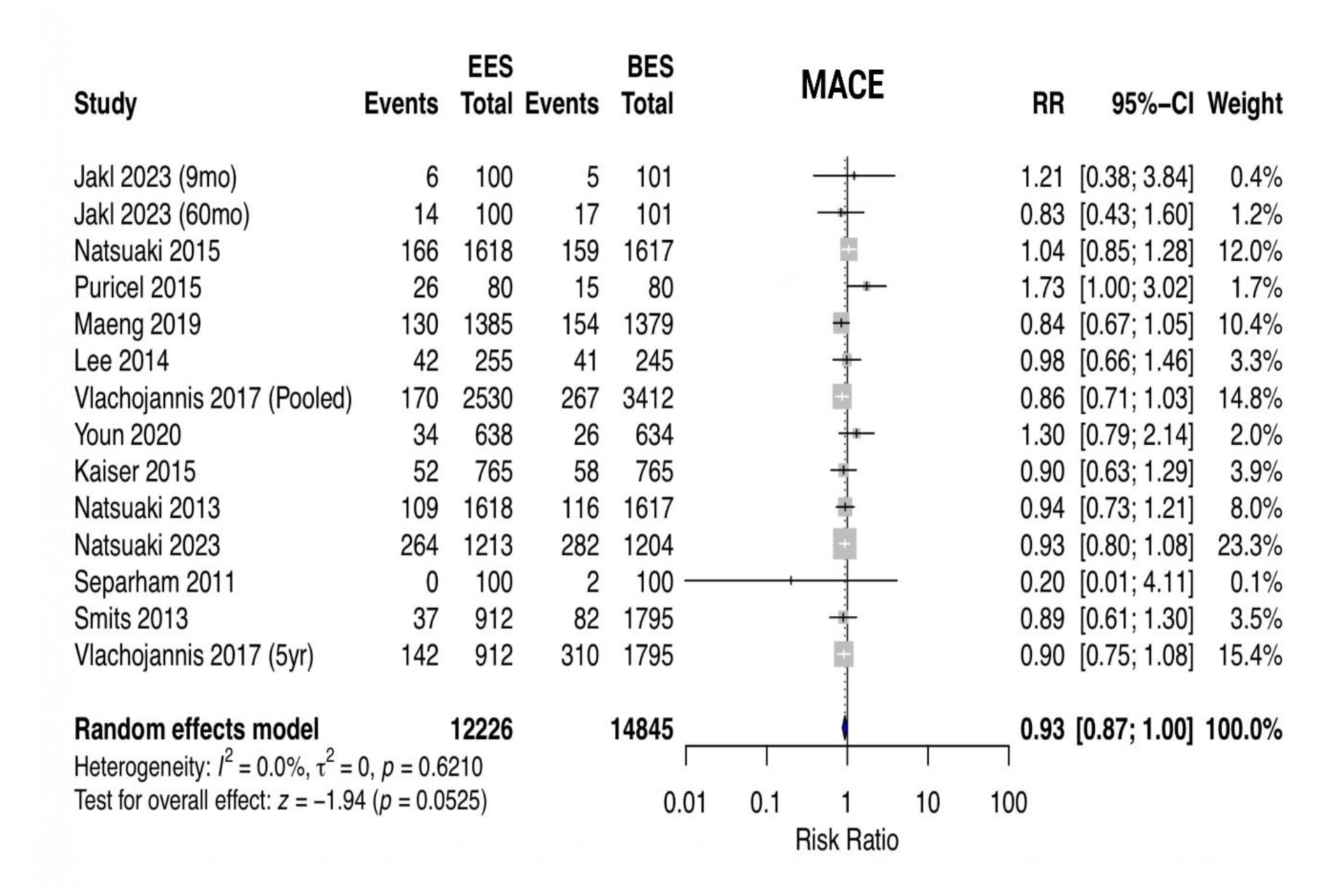
Forest Plot of Major Adverse Cardiac Events. Random-effects meta-analysis of major adverse cardiac events comparing everolimus-eluting stents (EES) with biolimus-eluting stents (BES). The forest plot displays individual study risk ratios with 95% confidence intervals (horizontal lines) and study weights (proportional to square size). The diamond represents the pooled risk ratio. Studies are ordered by publication year. CI, confidence interval; RR, risk ratio.

#### Device-Oriented Composite Endpoint (DOCE)

Ten trials with 24,939 patients reported DOCE. These studies similarly represented diverse patient populations with diabetes prevalence ranging from 19% to 46% and varying proportions of acute coronary syndrome presentations (16-100%). Risk of bias was predominantly low across contributing studies. The incidence was 9.4% (1,152/11,161) with EES versus 10.0% (1,479/13,778) with BES, yielding an RR of 0.94 (95% CI: 0.88-1.01; p=0.10), with no heterogeneity (I²=0%, p=0.59) (Supplementary Figure 2).

### Secondary Outcomes

#### All-Cause Mortality

Twelve trials (26,870 patients) with follow-up periods from 9 to 120 months contributed to this analysis. Study quality was high with consistent outcome ascertainment. Death occurred in 6.5% (795/12,126) of EES patients versus 6.5% (961/14,744) of BES patients (RR 0.96, 95% CI: 0.88-1.05; p=0.37; I²=0%) **(Figure 3A)**.

**Figure 3.**
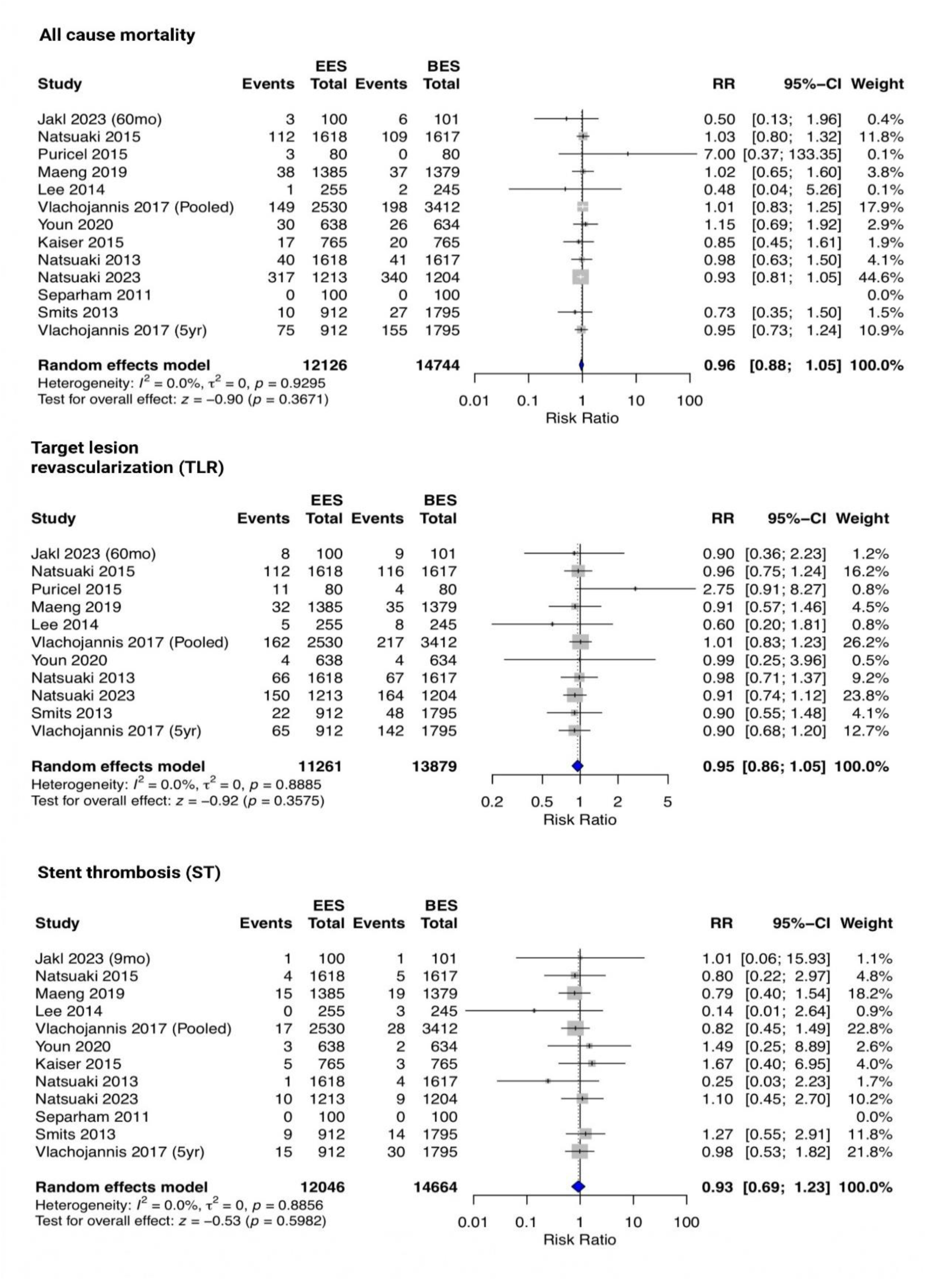
Forest Plots of Secondary Outcomes. Forest plots showing random-effects meta-analyses for (A) all-cause mortality, (B) target lesion revascularization, and (C) stent thrombosis comparing everolimus-eluting stents with biolimus-eluting stents-conventions as in Figure 2.

### Target Lesion Revascularization

Eleven trials (25,140 patients) reported clinically driven TLR using standardized definitions. All studies employed blinded clinical events committees. The incidence was 5.6% (637/11,261) with EES versus 5.8% (814/13,879) with BES (RR 0.95, 95% CI: 0.86-1.05; p=0.36; I²=0%) **(Figure 3B)**. The NNT was 370.3 (95% CI: 124 to ∞).

### Stent Thrombosis

Eleven trials (26,710 patients) reported definite or probable stent thrombosis according to Academic Research Consortium criteria, ensuring consistent case definitions across studies. The incidence was 0.7% (80/12,046) with EES versus 0.8% (118/14,664) with BES (RR 0.93, 95% CI: 0.69-1.23; p=0.60; I²=0%) **(Figure 3C)**. Analysis by timing revealed no significant differences in early (≤30 days: RR 1.05, 95% CI: 0.65-1.71), late (31 days-1 year: RR 0.84, 95% CI: 0.44-1.59), or very late (>1 year: RR 0.88, 95% CI: 0.54-1.44) stent thrombosis.

### Subgroup Analyses

Pre-specified subgroup analysis by follow-up duration showed consistent results across time periods **(Figure 4)**. For MACE, the RR was 0.95 (95% CI: 0.81-1.11) at ≤1 year, 0.96 (95% CI: 0.83-1.10) at 1-5 years, and 0.92 (95% CI: 0.82-1.02) at >5 years (p for interaction=0.88). Similar consistency was observed for DOCE across follow-up strata (p for interaction=0.89).

**Figure 4.**
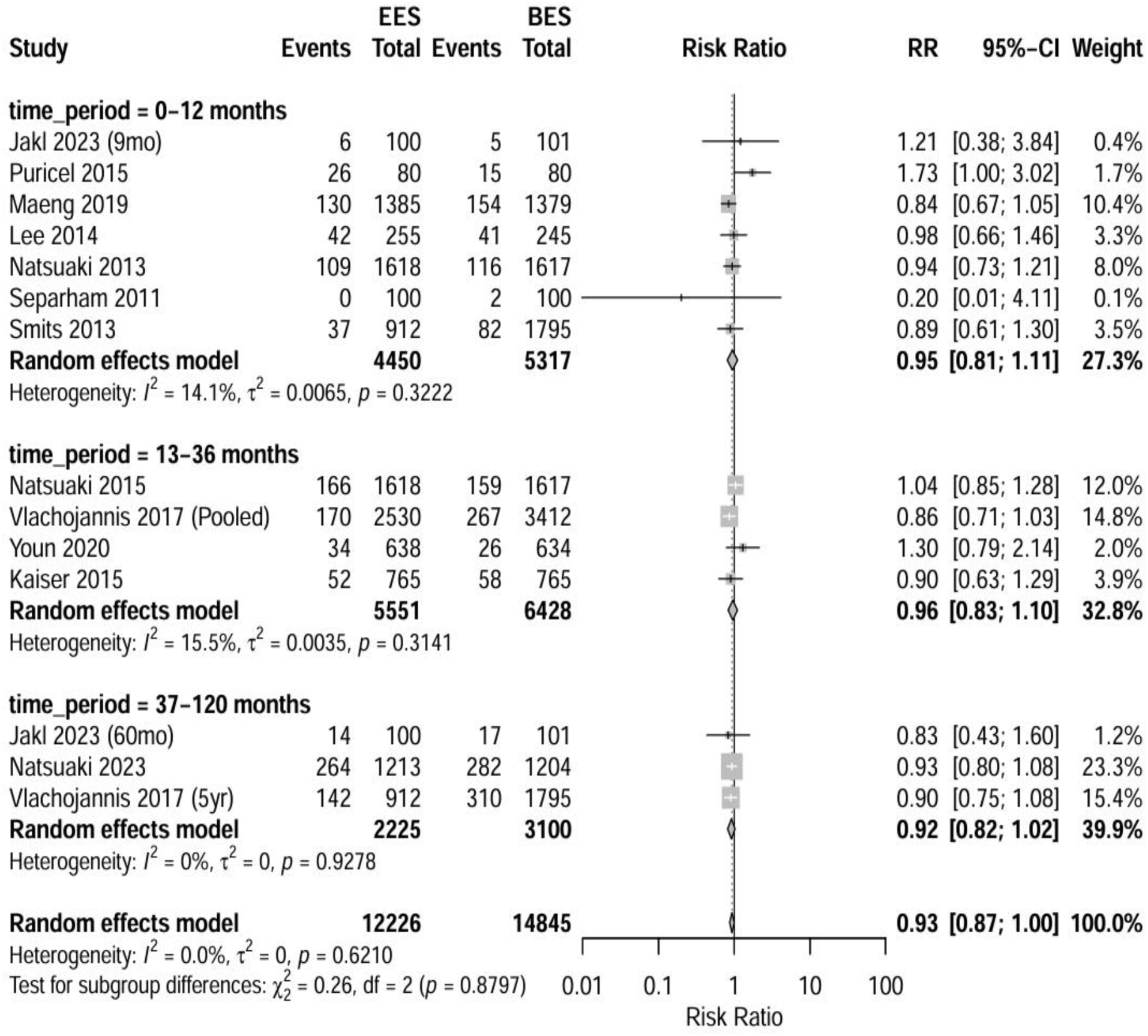
Subgroup Analysis by Follow-up Duration. Forest plot showing major adverse cardiac events stratified by follow-up duration: ≤1 year, 1-5 years, and >5 years. The test for subgroup differences showed no significant interaction (p=0.88), indicating a consistent treatment effect across time periods.

### Sensitivity Analyses

Leave-one-out analysis demonstrated the robustness of the primary outcome, with RR ranging from 0.92 to 0.95 across analyses (Supplementary Figure 3). Excluding trials with follow-up <1 year yielded an RR of 0.93 (95% CI: 0.87-1.00). Restricting analysis to trials with >1,000 patients (n=5 trials) showed an RR of 0.92 (95% CI: 0.85-0.99).

Cumulative meta-analysis ordered by publication year showed that the pooled effect estimate stabilized after inclusion of approximately 10,000 patients, with minimal change in subsequent years (Supplementary Figure 4).

### Publication Bias

Visual inspection of funnel plots for all outcomes showed symmetrical distribution **(Figure 5)**. Egger’s test revealed no evidence of small-study effects for MACE (p=0.74), mortality (p=0.82), TLR (p=0.68), or stent thrombosis (p=0.91). The trim-and-fill method identified no missing studies, with the adjusted estimate remaining unchanged.

**Figure 5.**
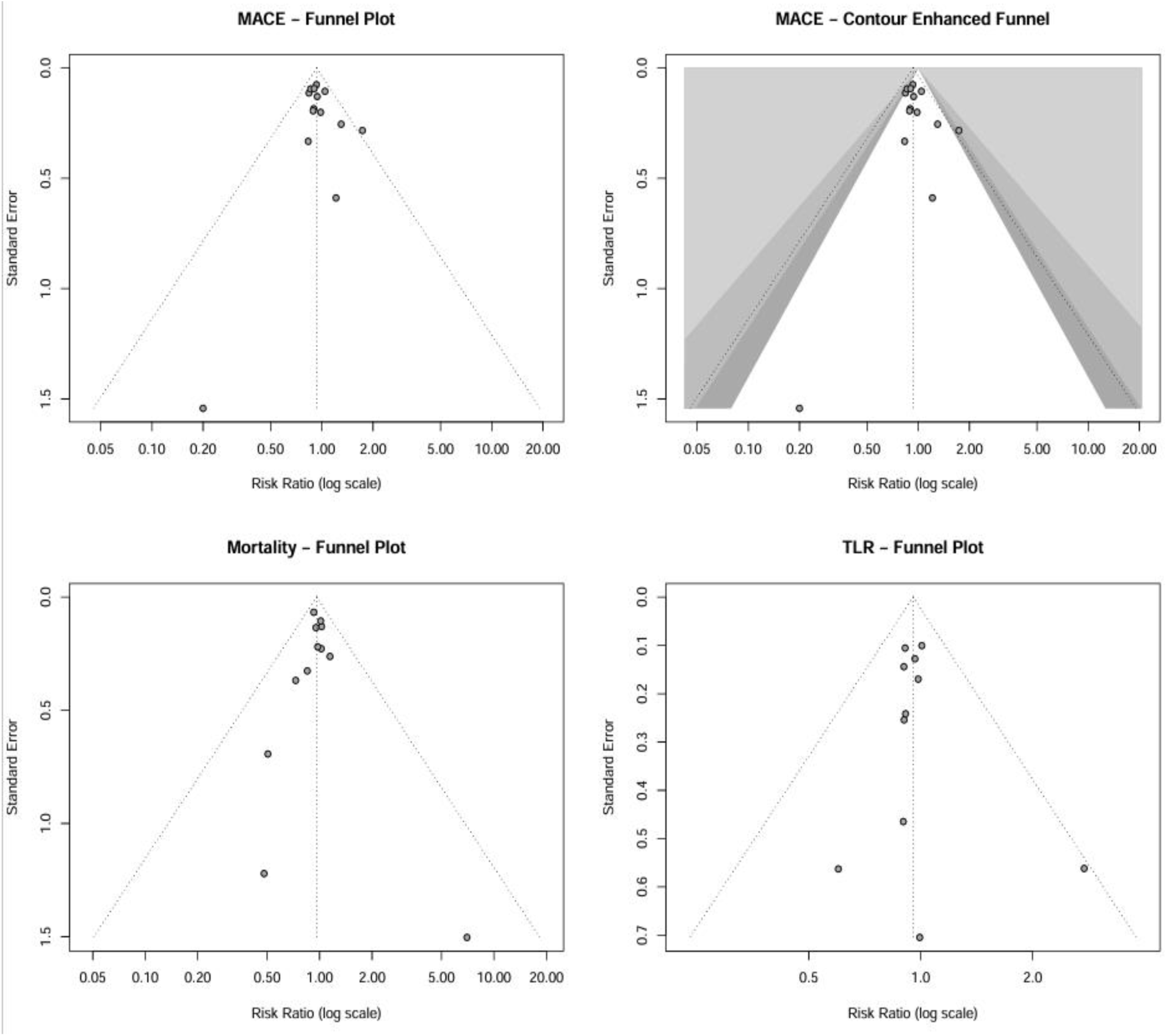
Funnel Plots for Assessment of Publication Bias. Funnel plots for (A) major adverse cardiac events, (B) all-cause mortality, and (C) target lesion revascularization. Each point represents a study, with the x-axis showing the log risk ratio and the y-axis showing the standard error. The vertical line indicates the pooled effect estimate. Symmetrical distribution suggests the absence of publication bias, confirmed by non-significant Egger’s tests.

### Meta-Regression Analyses

Univariable meta-regression analyses found no significant associations between treatment effect and study-level covariates **(Figure 6)**. The RR for MACE was not significantly influenced by mean follow-up duration (p=0.71), diabetes prevalence (p=0.48), proportion of ACS patients (p=0.82), or publication year (p=0.65). Similarly, the strut thickness difference between platforms did not significantly modify the treatment effect (p=0.34).

**Figure 6.**
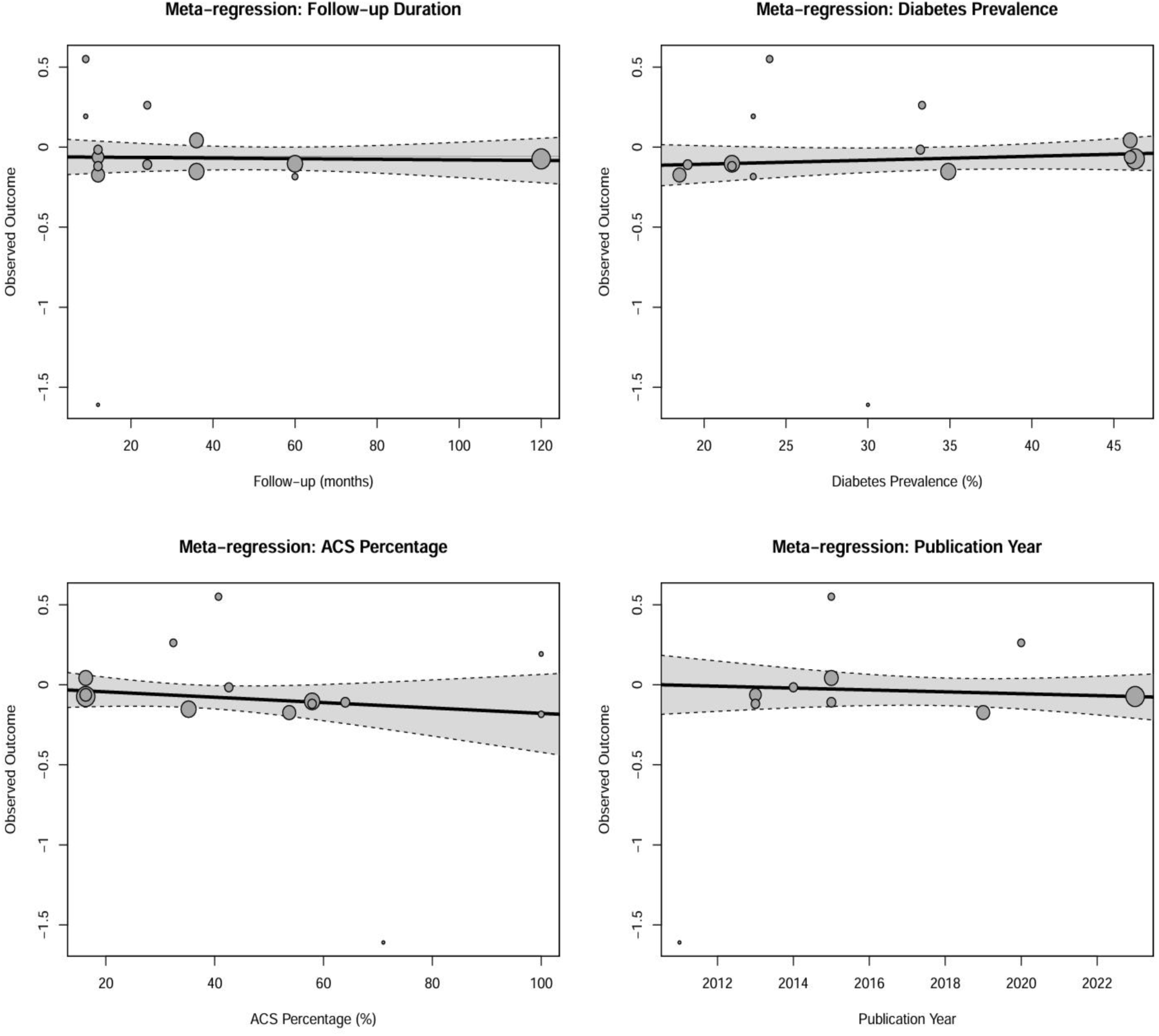
Meta-Regression Analyses. Bubble plots showing the relationship between treatment effect (log risk ratio for major adverse cardiac events) and study-level covariates: (A) follow-up duration, (B) diabetes prevalence, (C) proportion of patients with acute coronary syndromes, and (D) publication year. Circle size is proportional to study weight. The regression line with a 95% confidence interval shows no significant associations.

**Figure 7.**
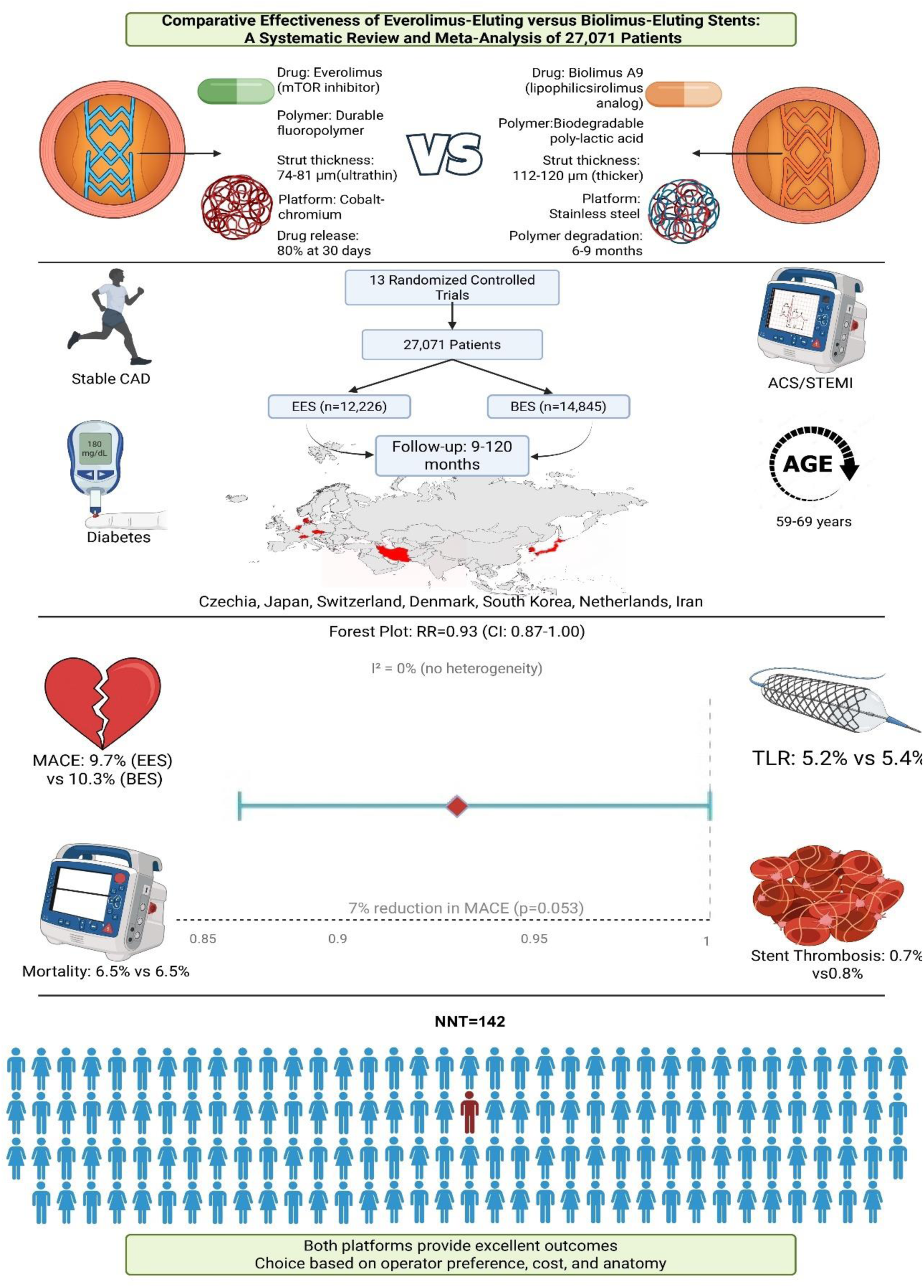
Central Illustration. This Central Illustration summarizes the key findings from this systematic review and meta-analysis comparing everolimus-eluting stents (EES) and biolimus-eluting stents (BES) in 27,071 patients. The graphic details the distinct pharmacological and structural properties of each stent platform and presents the primary clinical outcome, a non-significant 7% reduction in major adverse cardiovascular events (MACE) with EES (RR 0.93, 95% CI 0.87-1.00; p=0.053). The results demonstrate that both stent types provide excellent and comparable safety and efficacy profiles. Abbreviations: ACS, acute coronary syndrome; BES, biolimus-eluting stent; CAD, coronary artery disease; CI, confidence interval; EES, everolimus-eluting stent; MACE, major adverse cardiovascular events; mTOR, mammalian target of rapamycin; NNT, number needed to treat; RR, risk ratio; STEMI, ST-elevation myocardial infarction.

### Assessment of Reporting Bias

Assessment of selective outcome reporting revealed no evidence of reporting bias across the included studies. All trials reported their prespecified primary endpoints, and secondary outcomes were consistently reported. No trials showed evidence of post-hoc outcome selection or selective reporting of favorable results. The comprehensive reporting of both positive and neutral findings across all included trials supports the validity of our meta-analysis results.

### Quality of Evidence (GRADE)

The GRADE assessment indicated moderate certainty of evidence for MACE, all-cause mortality, and TLR, downgraded one level for serious risk of bias due to the open-label design of most trials **(Table 2)**. For stent thrombosis, the certainty was low, downgraded for both risk of bias and imprecision due to wide confidence intervals encompassing both benefit and harm.

**Table 2.**
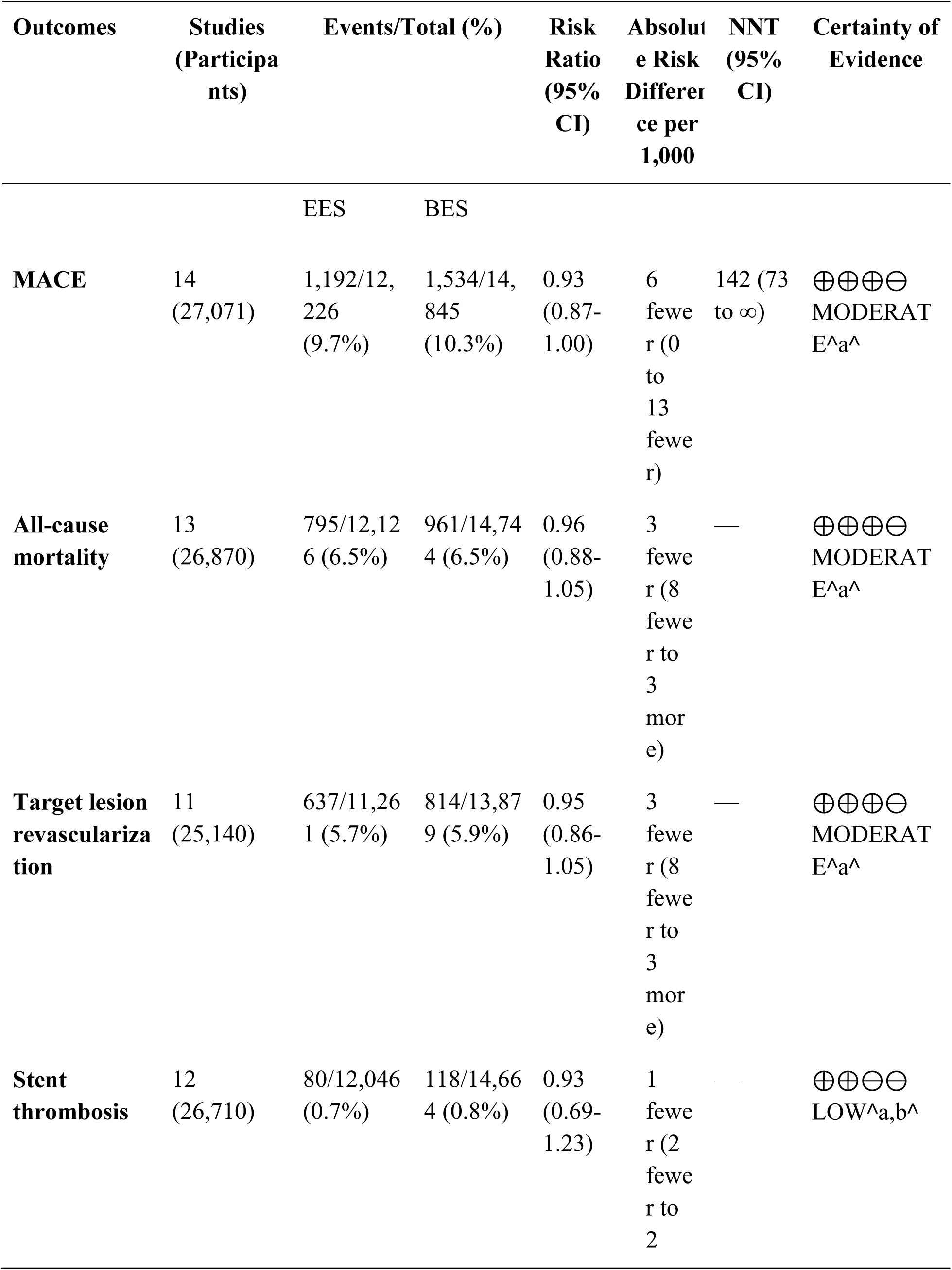
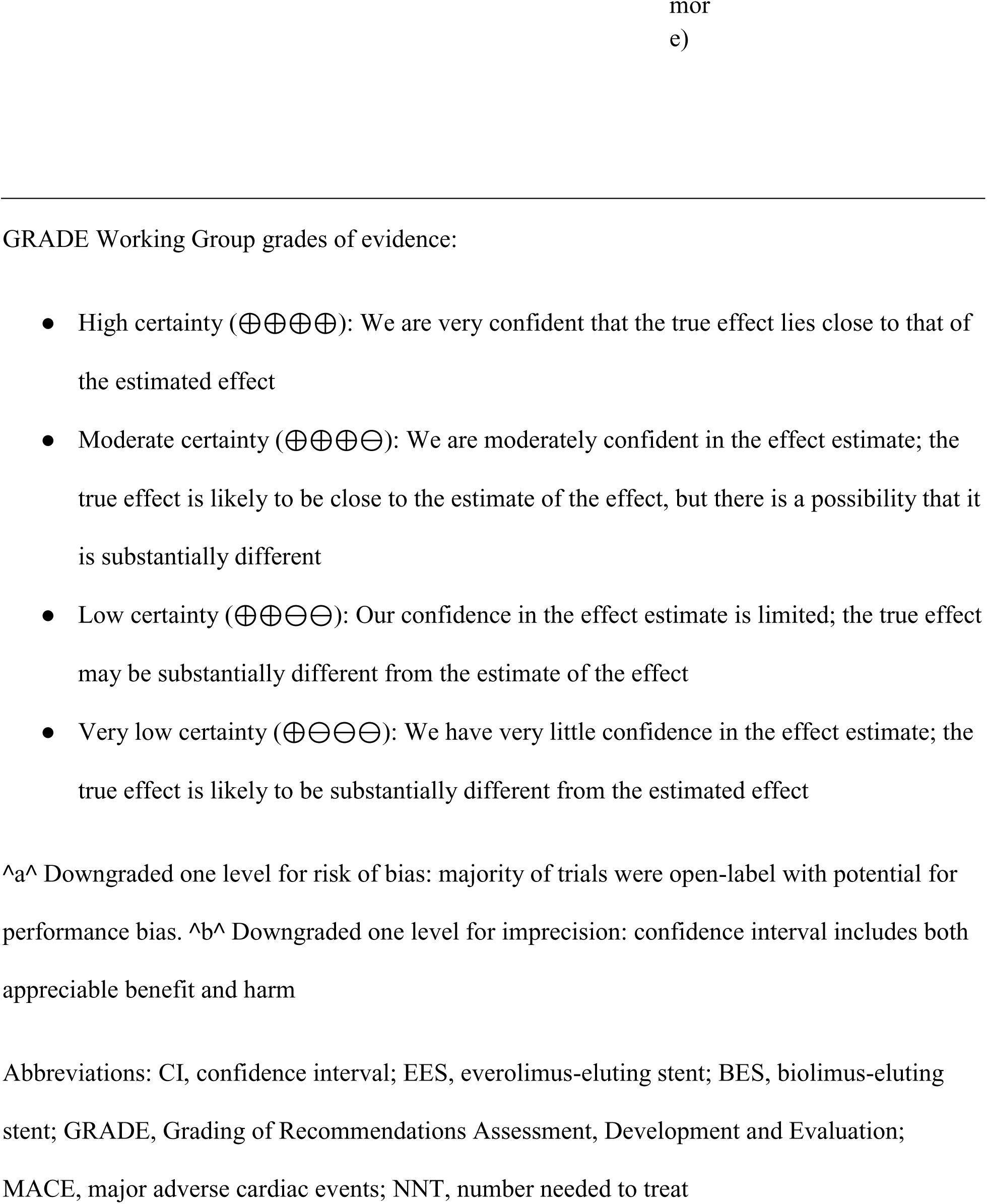
GRADE Summary of Findings: Everolimus-Eluting Stents Compared with Biolimus-Eluting Stents.

**Table 3.** Stent Platform Characteristics.

| **Characteristic** | **Everolimus-Eluting Stents** | **Biolimus-Eluting Stents** | **P value** |
| --- | --- | --- | --- |
| **Platform names** |  |  |  |
| Primary platforms | Xience/Promus (92%) | Nobori (62%), BioMatrix (38%) | — |
| Manufacturer | Abbott/Boston Scientific | Terumo/Biosensors | — |
| **Technical specifications** |  |  |  |
| Strut thickness (μm) | 74-81 | 112-120 | <0.001 |
| Polymer type | Durable fluoropolymer (92%) | Biodegradable poly-lactic acid (100%) | <0.001 |
| Drug dose (μg/cm<sup>2</sup>) | 100 | 15.6 | — |
| Drug release (% at 30 days) | 80% | 50% | — |
| **Procedural characteristics** |  |  |  |
| Number of stents/patient | 1.5 ± 0.6 | 1.5 ± 0.6 | 0.84 |
| Total stent length (mm) | 32.1 ± 18.0 | 32.7 ± 18.4 | 0.71 |
| Stent diameter (mm) | 3.0 ± 0.4 | 3.0 ± 0.4 | 0.92 |
| Direct stenting (%) | 18.4 | 17.9 | 0.68 |
| Post-dilatation (%) | 48.2 | 49.1 | 0.54 |

Values are mean ± SD or % unless otherwise indicated.

These GRADE assessments are summarized in **Table 2** and indicate that while the evidence is generally robust, the open-label nature of device trials introduces some uncertainty in the estimates of effect. No downgrades were applied for inconsistency (I²=0% for all outcomes), indirectness, or publication bias.

### Summary of Findings

In this comprehensive meta-analysis of 13 randomized trials including 27,071 patients, everolimus-eluting stents demonstrated a trend toward reduced MACE compared with biolimus-eluting stents (RR 0.93, 95% CI: 0.87-1.00; p=0.053), though this did not reach statistical significance. The GRADE assessment indicated moderate certainty of evidence for most outcomes, with consistent treatment effects across all secondary endpoints. The treatment effect was consistent across all secondary outcomes, subgroups, and sensitivity analyses, with remarkable homogeneity (I²=0%) suggesting true similarity in performance between these contemporary drug-eluting stent platforms.

## DISCUSSION

### Principal Findings

In this comprehensive meta-analysis of 13 randomized controlled trials including 27,071 patients, we found that everolimus-eluting stents were associated with a trend toward reduced major adverse cardiac events compared with biolimus-eluting stents (RR 0.93, 95% CI: 0.87-1.00; p=0.053). This 7% relative risk reduction approached but did not achieve conventional statistical significance. The absolute risk difference was modest at 0.7%, corresponding to a number needed to treat of 142 to prevent one MACE. Notably, we observed remarkable consistency across trials with no statistical heterogeneity (I²=0%), suggesting that these findings reflect true similarities in performance between contemporary drug-eluting stent platforms rather than statistical uncertainty.

### Comparison with Previous Evidence

Our findings align with and extend previous meta-analyses comparing biodegradable polymer DES with durable polymer DES. El-Hayek et al. analyzed 16 RCTs with 19,886 patients comparing biodegradable polymer DES (including BES) with second-generation durable polymer DES (including EES) [8], finding no significant differences in target vessel revascularization, cardiac death, or myocardial infarction. However, our analysis provides the first comprehensive direct comparison exclusively between EES and BES platforms, avoiding the heterogeneity introduced by mixing different drug types.

Sakurai et al. analyzed 8 RCTs involving 8,436 patients, with the most recent study published in 2016 and a maximum follow-up duration of 24 months [29]. However, they did not report on MACE or device-oriented composite endpoints (DOCE). In contrast, our study includes 4 additional RCTs published after 2016, totaling 5 RCTs with follow-up durations beyond 2 years, including 2 with 5-year and 1 with 10-year follow-up, allowing for a more comprehensive analysis of both MACE and DOCE.

The patient-level pooled analysis by Vlachojannis et al. of the NEXT and COMPARE II trials (5,942 patients) reported similar 3-year outcomes between BES and EES but noted a higher rate of target vessel myocardial infarction with BES [30]. Our broader analysis, incorporating 11 additional trials with longer follow-up, suggests this difference may not persist when examining the totality of evidence. Importantly, the network meta-analysis by Palmerini et al. suggested that cobalt-chromium EES was associated with lower mortality and stent thrombosis compared with BES [9], findings that our direct comparison supports, albeit without reaching statistical significance.

### Mechanistic Considerations

The observed trend favoring EES may be explained by fundamental differences in stent design. First, strut thickness differs substantially between platforms, with EES utilizing ultrathin struts (74-81 μm) compared with thicker BES struts (112-120 μm). The meta-analysis by Bangalore et al. demonstrated that ultrathin strut DES (<80 μm) were associated with a 16% reduction in target lesion failure compared with thicker strut DES [7]. Thinner struts improve arterial healing, reduce flow disturbance, and minimize vessel injury [2].

Second, the polymer technology differs fundamentally between platforms. While the theoretical advantages of biodegradable polymers include reduced late inflammatory responses and potential for complete vessel healing [4], our analysis suggests these benefits may not translate into superior clinical outcomes within the 10-year follow-up window. The durable fluoropolymer used in EES has demonstrated excellent biocompatibility with minimal inflammatory response [2], potentially explaining the comparable or slightly superior outcomes despite permanent polymer presence.

### Clinical Implications

Our findings have important implications for clinical practice. Both EES and BES demonstrate excellent safety and efficacy profiles, with MACE rates of approximately 10% at a mean follow-up of 3 years. The absolute difference between platforms is small (0.7%), suggesting that stent selection may reasonably be based on other considerations, including cost, availability, operator experience, and specific anatomical considerations.

The absence of heterogeneity across diverse patient populations (diabetes prevalence 19-46%, ACS presentation 16-100%) and follow-up durations (9-120 months) suggests these findings are broadly applicable. The consistency of results in leave-one-out analysis further supports the generalizability of our conclusions. Notably, neither platform demonstrated superiority in preventing stent thrombosis, with very low rates (<1%) observed with both EES and BES, confirming the safety of contemporary DES technology when combined with current antiplatelet therapy protocols [13].

### Strengths and Limitations

This meta-analysis has several strengths. First, we included only direct randomized comparisons between EES and BES, avoiding the assumptions required for network meta-analyses. Second, the large sample size (27,071 patients) provided adequate statistical power to detect clinically meaningful differences. Third, the absence of statistical heterogeneity and publication bias, confirmed through multiple methods, strengthens confidence in our findings. Fourth, the inclusion of long-term follow-up data, with trials reporting outcomes beyond 5 years, addresses concerns about very late events with different polymer technologies.

However, important limitations merit consideration. First, the open-label design of most included trials introduces potential performance bias, though this was mitigated by blinded endpoint adjudication committees. This limitation led to downgrading the certainty of evidence to moderate for most outcomes in our GRADE assessment. Second, we analyzed aggregate rather than individual patient data, precluding examination of treatment effects in specific patient subgroups that might benefit differentially from either platform. Third, the included trials used various definitions for composite endpoints, though this did not introduce statistical heterogeneity. Fourth, technological iterations within each platform family over the study period may have influenced outcomes, though our meta-regression found no significant temporal trends. Fifth, bleeding outcomes could not be analyzed due to inconsistent reporting across trials. Additionally, while DAPT duration ranged from 6-12 months, detailed protocols were not available for subgroup analysis. Sixth, BES platforms have limited current commercial availability in some markets, which may affect the generalizability of our findings to contemporary practice. Finally, variations in dual antiplatelet therapy protocols across trials may have influenced outcomes, though current guidelines do not differentiate recommendations based on polymer type.

### Future Directions

Several areas warrant future investigation. First, very long-term follow-up beyond 10 years will be important to determine whether biodegradable polymers confer late benefits after complete polymer degradation. The ongoing extended follow-up of trials like NEXT [31] will provide valuable insights. Second, comparison of the newest-generation platforms, including biodegradable polymer EES (Synergy) versus thin-strut BES, would address whether technological advances have altered the comparative effectiveness. Third, individual patient data meta-analysis could identify specific subgroups who might benefit preferentially from either platform, enabling personalized stent selection. Fourth, cost-effectiveness analyses incorporating the small absolute differences in clinical outcomes would inform healthcare policy decisions. Fifth, future trials should adopt standardized bleeding definitions to enable comprehensive safety comparisons between stent platforms.

The development of novel technologies, including bioresorbable scaffolds, polymer-free drug-eluting stents, and drug-coated balloons, may ultimately supersede current-generation metallic DES. However, our analysis confirms that both contemporary EES and BES platforms provide excellent clinical outcomes, setting a high benchmark for future innovations.

## CONCLUSIONS

In this comprehensive meta-analysis of randomized trials, everolimus-eluting stents demonstrated a trend toward reduced major adverse cardiac events compared with biolimus-eluting stents, though this did not reach statistical significance. The remarkable consistency of findings across diverse populations and extended follow-up periods suggests both platforms provide excellent clinical outcomes with subtle differences that may be attributed to strut thickness and polymer technology. Given the small absolute differences and moderate certainty of evidence, stent selection between these contemporary platforms may reasonably be individualized based on operator preference, cost considerations, and specific clinical scenarios. These findings support the continued use of both EES and BES as first-line options for percutaneous coronary intervention while awaiting longer-term data and next-generation technologies.

## Data Availability

All data generated or analyzed during this study are included in this published article and its supplementary information files. The datasets extracted from the included studies are available from the corresponding author upon reasonable request.

## Acknowledgements

We thank all the study participants.

## Disclosures

### Ethics approval and consent to participate

Ethical approval was not required for this meta-analysis as it involved the analysis of previously published randomized controlled trials with publicly available data. No primary data collection involving human participants was conducted as part of this study.

### Consent for publication

Not applicable.

### Availability of data and materials

The extracted data supporting the findings of this meta-analysis, including event counts and effect estimates for all included studies, are available in this published article and its supplementary files. The analysis code (R scripts) used for all meta-analyses is available from the corresponding author upon reasonable request. Primary data remain with the original trial investigators and are subject to their respective data sharing policies. The PRISMA checklist and completed PROSPERO registration (CRD42025108092) are publicly available.

## Authors’ Contributions

Conceptualization and Study Design: IMK, SB, EK, AK

Data Collection and Investigation: VB, SSN, MC, JJP, ZAS, YPB, HUW, SA, OT, JMC, MSK

Data Analysis and Interpretation: IMK, SB, EK, AK,

Manuscript Writing - Original Draft: IMK, SB, EK

Manuscript Writing - Review and Editing: All authors

Supervision: IMK, EK, MSK

Project Administration: IMK, AK

Critical Revision of the Manuscript: EK, AK

Final Approval of the Manuscript: All authors

All authors have read and agreed to the published version of the manuscript.

## Non-standard Abbreviations and Acronyms

ACS: Acute coronary syndrome
BES: Biolimus-eluting stents
CI: Confidence interval
DES: Drug-eluting stents
DOCE: Device-oriented composite endpoint
EES: Everolimus-eluting stents
MACE: Major adverse cardiac events
NNT: Number needed to treat
RCT: Randomized controlled trial
RR: Risk ratio
STEMI: ST-elevation myocardial infarction
TLR: Target lesion revascularization

## REFERENCES

1. Writing Committee Members, Lawton JS, Tamis-Holland JE, Bangalore S, Bates ER, Beckie TM, Bischoff JM, Bittl JA, Cohen MG, DiMaio JM, Don CW. 2021 ACC/AHA/SCAI guideline for coronary artery revascularization: a report of the American College of Cardiology/American Heart Association Joint Committee on Clinical Practice Guidelines. Journal of the American College of Cardiology. 2022 Jan 18;79(2):e21–129. doi:10.1016/j.jacc.2021.09.006

2. Torii S, Jinnouchi H, Sakamoto A, Kutyna M, Cornelissen A, Kuntz S, Guo L, Mori H, Harari E, Paek KH, Fernandez R. Drug-eluting coronary stents: insights from preclinical and pathology studies. Nature Reviews Cardiology. 2020 Jan;17(1):37–51. doi:10.1038/s41569-019-0234-x

3. Byrne RA, Serruys PW, Baumbach A, Escaned J, Fajadet J, James S, Joner M, Oktay S, Jüni P, Kastrati A, Sianos G. Report of a European Society of Cardiology-European Association of Percutaneous Cardiovascular Interventions task force on the evaluation of coronary stents in Europe: executive summary. European heart journal. 2015 Oct 7;36(38):2608–20. doi:10.1093/eurheartj/ehv203

4. Stefanini GG, Holmes Jr DR. Drug-eluting coronary-artery stents. New England Journal of Medicine. 2013 Jan 17;368(3):254–65. doi:10.1056/NEJMra1210816

5. Windecker S, Serruys PW, Wandel S, Buszman P, Trznadel S, Linke A, Lenk K, Ischinger T, Klauss V, Eberli F, Corti R. Biolimus-eluting stent with biodegradable polymer versus sirolimus-eluting stent with durable polymer for coronary revascularisation (LEADERS): a randomised non-inferiority trial. The Lancet. 2008 Sep 27;372(9644):1163-73. . doi:10.1016/S0140-6736(08)61244-1

6. Bangalore S, Toklu B, Patel N, Feit F, Stone GW. Newer-generation ultrathin strut drug-eluting stents versus older second-generation thicker strut drug-eluting stents for coronary artery disease: meta-analysis of randomized trials. Circulation. 2018 Nov 13;138(20):2216–26. 118.034456

7. Bangalore S, Toklu B, Amoroso N, Fusaro M, Kumar S, Hannan EL, Faxon DP, Feit F. Bare metal stents, durable polymer drug eluting stents, and biodegradable polymer drug eluting stents for coronary artery disease: mixed treatment comparison meta-analysis. Bmj. 2013 Nov 8;347. doi:10.1136/bmj.f6625

8. El-Hayek G, Bangalore S, Casso Dominguez A, Devireddy C, Jaber W, Kumar G, Mavromatis K, Tamis-Holland J, Samady H. Meta-analysis of randomized clinical trials comparing biodegradable polymer drug-eluting stent to second-generation durable polymer drug-eluting stents. JACC: Cardiovascular Interventions. 2017 Mar 13;10(5):462–73. doi:10.1016/j.jcin.2016.12.002

9. Palmerini T, Biondi-Zoccai G, Della Riva D, Mariani A, Sabaté M, Smits PC, Kaiser C, D’Ascenzo F, Frati G, Mancone M, Genereux P. Clinical outcomes with bioabsorbable polymer-versus durable polymer-based drug-eluting and bare-metal stents: evidence from a comprehensive network meta-analysis. Journal of the American College of Cardiology. 2014 Feb 4;63(4):299–307. doi:10.1016/j.jacc.2013.09.061

10. Natsuaki M, Kozuma K, Morimoto T, Kadota K, Muramatsu T, Nakagawa Y, Akasaka T, Igarashi K, Tanabe K, Morino Y, Ishikawa T. Biodegradable polymer biolimus-eluting stent versus durable polymer everolimus-eluting stent: a randomized, controlled, noninferiority trial. Journal of the American College of Cardiology. 2013 Jul 16;62(3):181–90. doi:10.1016/j.jacc.2013.04.045

11. Smits PC, Hofma S, Togni M, Vázquez N, Valdés M, Voudris V, Slagboom T, Goy JJ, Vuillomenet A, Serra A, Nouche RT. Abluminal biodegradable polymer biolimus-eluting stent versus durable polymer everolimus-eluting stent (COMPARE II): a randomised, controlled, non-inferiority trial. The Lancet. 2013 Feb 23;381(9867):651–60. doi:10.1016/S0140-6736(12)61852-2

12. Vlachojannis GJ, Smits PC, Hofma SH, Togni M, Vázquez N, Valdés M, Voudris V, Slagboom T, Goy JJ, den Heijer P, van der Ent M. Biodegradable polymer biolimus-eluting stents versus durable polymer everolimus-eluting stents in patients with coronary artery disease: final 5-year report from the COMPARE II trial (abluminal biodegradable polymer biolimus-eluting stent versus durable polymer everolimus-eluting stent). Cardiovascular Interventions. 2017 Jun 26;10(12):1215–21. doi:10.1016/j.jcin.2017.02.029

13. Valgimigli M, Bueno H, Byrne RA, Collet JP, Costa F, Jeppsson A, Jüni P, Kastrati A, Kolh P, Mauri L, Montalescot G. 2017 ESC focused update on dual antiplatelet therapy in coronary artery disease developed in collaboration with EACTS: The Task Force for dual antiplatelet therapy in coronary artery disease of the European Society of Cardiology (ESC) and of the European Association for Cardio-Thoracic Surgery (EACTS). European heart journal. 2018 Jan 14;39(3):213–60. doi:10.1093/eurheartj/ehx419

14. Neumann FJ, Sousa-Uva M, Ahlsson A, Alfonso F, Banning AP, Benedetto U, Byrne RA, Collet JP, Falk V, Head SJ, Jüni P. 2018 ESC/EACTS Guidelines on myocardial revascularization. EuroIntervention. 2019 Feb 20;14(14):1435–534. doi:10.4244/EIJY19M01_01

15. Moher D, Shamseer L, Clarke M, Ghersi D, Liberati A, Petticrew M, Shekelle P, Stewart LA, Prisma-P Group. Preferred reporting items for systematic review and meta-analysis protocols (PRISMA-P) 2015 statement. Systematic reviews. 2015 Jan 1;4(1):1. doi:10.1186/2046-4053-4-1

16. Page MJ, McKenzie JE, Bossuyt PM, Boutron I, Hoffmann TC, Mulrow CD, Shamseer L, Tetzlaff JM, Akl EA, Brennan SE, Chou R. The PRISMA 2020 statement: an updated guideline for reporting systematic reviews. bmj. 2021 Mar 29;372. doi:10.1136/bmj.n71

17. Garcia-Garcia HM, McFadden EP, Farb A, Mehran R, Stone GW, Spertus J, Onuma Y, Morel MA, van Es GA, Zuckerman B, Fearon WF. Standardized end point definitions for coronary intervention trials: the academic research consortium-2 consensus document. European heart journal. 2018 Jun 14;39(23):2192–207. doi:10.1093/eurheartj/ehy223

18. Higgins JP. Cochrane handbook for systematic reviews of interventions. Cochrane Collaboration and John Wiley & Sons Ltd. 2008.

19. Sterne JA, Savović J, Page MJ, Elbers RG, Blencowe NS, Boutron I, Cates CJ, Cheng HY, Corbett MS, Eldridge SM, Emberson JR. RoB 2: a revised tool for assessing risk of bias in randomised trials. bmj. 2019 Aug 28;366.

20. DerSimonian R, Laird N. Meta-analysis in clinical trials. Controlled clinical trials. 1986 Sep 1;7(3):177–88. doi:10.1016/0197-2456(86)90046-2

21. R Core Team (2024). R: A language and environment for statistical computing. R Foundation for Statistical Computing, Vienna, Austria. URL https://www.R-project.org/.

22. Higgins JP, Thompson SG, Deeks JJ, Altman DG. Measuring inconsistency in meta-analyses. bmj. 2003 Sep 4;327(7414):557–60.

23. Viechtbauer W, Cheung MW. Outlier and influence diagnostics for meta-analysis. Research synthesis methods. 2010 Apr;1(2):112–25.

24. Lau J, Antman EM, Jimenez-Silva J, Kupelnick B, Mosteller F, Chalmers TC. Cumulative meta-analysis of therapeutic trials for myocardial infarction. New England Journal of Medicine. 1992 Jul 23;327(4):248–54. doi:10.1056/NEJM199207233270406

25. Duval S, Tweedie R. Trim and fill: a simple funnel-plot–based method of testing and adjusting for publication bias in meta-analysis. Biometrics. 2000 Jun;56(2):455–63. doi:10.1111/j.0006-341X.2000.00455.x

26. Jakl M, Cervinka P, Kanovsky J, Kala P, Poloczek M, Cervinkova M, Bezerra HG, Valenta Z, Costa MA. Randomized comparison of 9-month stent strut coverage of biolimus and everolimus drug-eluting stents assessed by optical coherence tomography in patients with ST-segment elevation myocardial infarction. Long-term (5-years) clinical follow-up (ROBUST trial). Cardiology Journal. 2023;30(6):921–8. doi:10.5603/CJ.a2023.0013

27. Natsuaki M, Kozuma K, Morimoto T, Kadota K, Muramatsu T, Nakagawa Y, Akasaka T, Igarashi K, Tanabe K, Morino Y, Ishikawa T. Final 3-year outcome of a randomized trial comparing second-generation drug-eluting stents using either biodegradable polymer or durable polymer: NOBORI biolimus-eluting versus XIENCE/PROMUS everolimus-eluting stent trial. Circulation: Cardiovascular Interventions. 2015 Oct;8(10):e002817. doi:10.1161/CIRCINTERVENTIONS.115.002817

28. Lee JY, Park DW, Kim YH, Ahn JM, Kim WJ, Kang SJ, Lee SW, Lee CW, Park SW, Yun SC, Yang TH. Comparison of Biolimus A9–Eluting (Nobori) and Everolimus-Eluting (Promus Element) Stents in Patients With De Novo Native Long Coronary Artery Lesions: A Randomized Long Drug-Eluting Stent V Trial. Circulation: Cardiovascular Interventions. 2014 Jun;7(3):322–9. doi:10.1161/CIRCINTERVENTIONS.113.000841

29. Sakurai R, Burazor I, Bonneau HN, Kaneda H. Long-term outcomes of biodegradable polymer biolimus-eluting stents versus durable polymer everolimus-eluting stents: a meta-analysis of randomized controlled trials. International journal of cardiology. 2016 Nov 15;223:1066–71. doi:10.1016/j.ijcard.2016.07.186

30. Vlachojannis GJ, Puricel S, Natsuaki M, Morimoto T, Smits PC, Kimura T. Biolimus-eluting versus everolimus-eluting stents in coronary artery disease: a pooled analysis from the NEXT (NOBORI biolimus-eluting versus XIENCE/PROMUS everolimus-eluting stent) and COMPARE II (Abluminal biodegradable polymer biolimus-eluting stent versus durable polymer everolimus-eluting stent) randomised trials. EuroIntervention: journal of EuroPCR in collaboration with the Working Group on Interventional Cardiology of the European Society of Cardiology. 2017 Mar 1;12(16):1970–7. doi:10.4244/EIJ-D-16-00773

31. Natsuaki M, Watanabe H, Morimoto T, Kozuma K, Kadota K, Muramatsu T, Nakagawa Y, Akasaka T, Hanaoka KI, Tanabe K, Morino Y. Biodegradable or durable polymer drug-eluting stents in patients with coronary artery disease: ten-year outcomes of the randomised NEXT Trial. EuroIntervention. 2023 Aug 7;19(5):e402. doi:10.4244/EIJ-D-23-00076

32. Maeng M, Christiansen EH, Raungaard B, Kahlert J, Terkelsen CJ, Kristensen SD, Carstensen S, Aarøe J, Jensen SE, Villadsen AB, Lassen JF. Everolimus-eluting versus biolimus-eluting stents with biodegradable polymers in unselected patients undergoing percutaneous coronary intervention: a randomized noninferiority trial with 1-year follow-up (SORT OUT VIII Trial). JACC: Cardiovascular Interventions. 2019 Apr 8;12(7):624–33. doi:10.1016/j.jcin.2018.12.036

33. Puricel S, Arroyo D, Corpataux N, Baeriswyl G, Lehmann S, Kallinikou Z, Muller O, Allard L, Stauffer JC, Togni M, Goy JJ. Comparison of everolimus-and biolimus-eluting coronary stents with everolimus-eluting bioresorbable vascular scaffolds. Journal of the American College of Cardiology. 2015 Mar 3;65(8):791–801. doi:10.1016/j.jacc.2014.12.017

34. Kaiser C, Galatius S, Jeger R, Gilgen N, Jensen JS, Naber C, Alber H, Wanitschek M, Eberli F, Kurz DJ, Pedrazzini G. Long-term efficacy and safety of biodegradable-polymer biolimus-eluting stents: main results of the Basel Stent Kosten-Effektivitäts Trial–PROspective Validation Examination II (BASKET-PROVE II), a randomized, controlled noninferiority 2-year outcome trial. Circulation. 2015 Jan 6;131(1):74–81. doi:10.1161/CIRCULATIONAHA.114.013520

35. Youn YJ, Lee JW, Ahn SG, Lee SH, Yoon J, Park KS, Lee JB, Yoo SY, Lim DS, Cho JH, Choi CU. Randomized comparison of everolimus-and zotarolimus-eluting coronary stents with biolimus-eluting stents in all-comer patients. Circulation: Cardiovascular Interventions. 2020 Mar;13(3):e008525. doi:10.1161/CIRCINTERVENTIONS.119.008525

36. Separham A, Sohrabi B, Aslanabadi N, Ghaffari S. The twelve-month outcome of biolimus eluting stent with biodegradable polymer compared with an everolimus eluting stent with durable polymer. Journal of cardiovascular and thoracic research. 2011 Dec 28;3(4):113. doi:10.5681/jcvtr.2011.025

