## Supplementary Table S1, Supplementary Table S2., Supplementary Figure 1, Supplementary Figure 2., Supplementary Figure 3., Supplementary Figure 4. for "Comparative Clinical Outcomes of Everolimus versus Biolimus-Eluting Stents: A Meta-Analysis of 27,071 Patients from Randomized Trials"

**Supplementary Materials**

**Supplementary Table S1. Detailed Search Strategy**

| Database | Search string | Results | Date |
|---|---|---|---|
| **PubMed (MEDLINE)** | ((everolimus[tiab] OR "everolimus eluting"[tiab] OR EES[tiab] OR Xience[tiab] OR Promus[tiab] OR Synergy[tiab]) AND (biolimus[tiab] OR "biolimus eluting"[tiab] OR 7BES[tiab] OR Nobori[tiab] OR Biomatrix[tiab]) AND (randomized[tiab] OR randomised[tiab] OR RCT[tiab] OR "controlled trial"[tiab] OR "clinical trial"[pt] OR "randomized controlled trial"[pt]) AND (stent*[tiab] OR "percutaneous coronary"[tiab] OR PCI[tiab] OR "coronary intervention"[tiab])) | 84 | 25 May, 2025 |
| **Cochrane CENTRAL** | (everolimus OR "everolimus eluting" OR EES OR Xience OR Promus OR Synergy) AND (biolimus OR "biolimus eluting" OR BES OR Nobori OR Biomatrix) AND (stent* OR "percutaneous coronary" OR PCI OR "coronary intervention") | 163 | 25 May, 2025 |
| **Embase by Ovid** | ((everolimus or everolimus eluting or EES or Xience or Promus or Synergy) and (biolimus or biolimus eluting or BES or Nobori or Biomatrix) and (random$ or RCT or controlled trial or clinical trial) and (stent$ or percutaneous coronary or PCI or coronary intervention)).mp. | 587 | 25 May, 2025 |
| **Scopus** | TITLE-ABS-KEY((everolimus OR "everolimus eluting" OR EES OR Xience OR Promus OR Synergy) AND (biolimus OR "biolimus eluting" OR BES OR Nobori OR Biomatrix) AND (random* OR RCT OR "controlled trial" OR "clinical trial") AND (stent* OR "percutaneous coronary" OR PCI OR "coronary intervention")) | 421 | 25 May, 2025 |

**Supplementary Table S2. Full-Text Articles Assessed and Excluded (n=8)**

| Study | Design | Sample Size | Groups Compared | Follow-up | Reason for Exclusion |
|---|---|---|---|---|---|
| **COBRA II Trial** | RCT | 15 patients | • Axxess BES + Absorb BVS (n=8)<br>• Mod-T technique with Absorb BVS only (n=7) | 5 years | **No direct EES vs BES comparison**<br>• Compares bifurcation stenting strategies<br>• BES (Axxess) used in combination with EES scaffolds, not as standalone comparator<br>• Focus on bioresorbable scaffold technology |
| **ISAR-TEST-4 Trial** | RCT | 2,603 patients | •Biodegradable polymer rapamycin-eluting stent (n=1,299)<br>• Permanent polymer DES (n=1,304):- Cypher (sirolimus) n=652 - Xience (everolimus) n=652 | 12 months | **No biolimus-eluting stent included**<br>•Biodegradable polymer stent uses rapamycin/sirolimus, NOT biolimus<br>• Does not meet inclusion criterion of EES vs BES comparison |
| **Gyldenkerne 2019** | Subgroup analysis | Not specified | EES vs BES in:<br>• Patients with diabetes | Not specified | **Subgroup comparison only**<br>• Not a primary RCT |

| | | | | | |
|---|---|---|---|---|---|
| | | | • Patients without diabetes | | • Analyzed specific patient subgroups from existing trial data |
| **Conference Papers** | Various | Various | Various | Various | **Publication type** • Conference abstracts/papers (n=5) • Not full peer-reviewed publications |

**Summary of Exclusions**

| Exclusion Reason | Number of Studies |
|---|---|
| No direct EES vs BES comparison | 2 |
| Subgroup analysis | 1 |
| Conference papers | 5 |
| **Total Excluded** | **8** |

**Supplementary Figure 1. Risk of Bias Assessment**

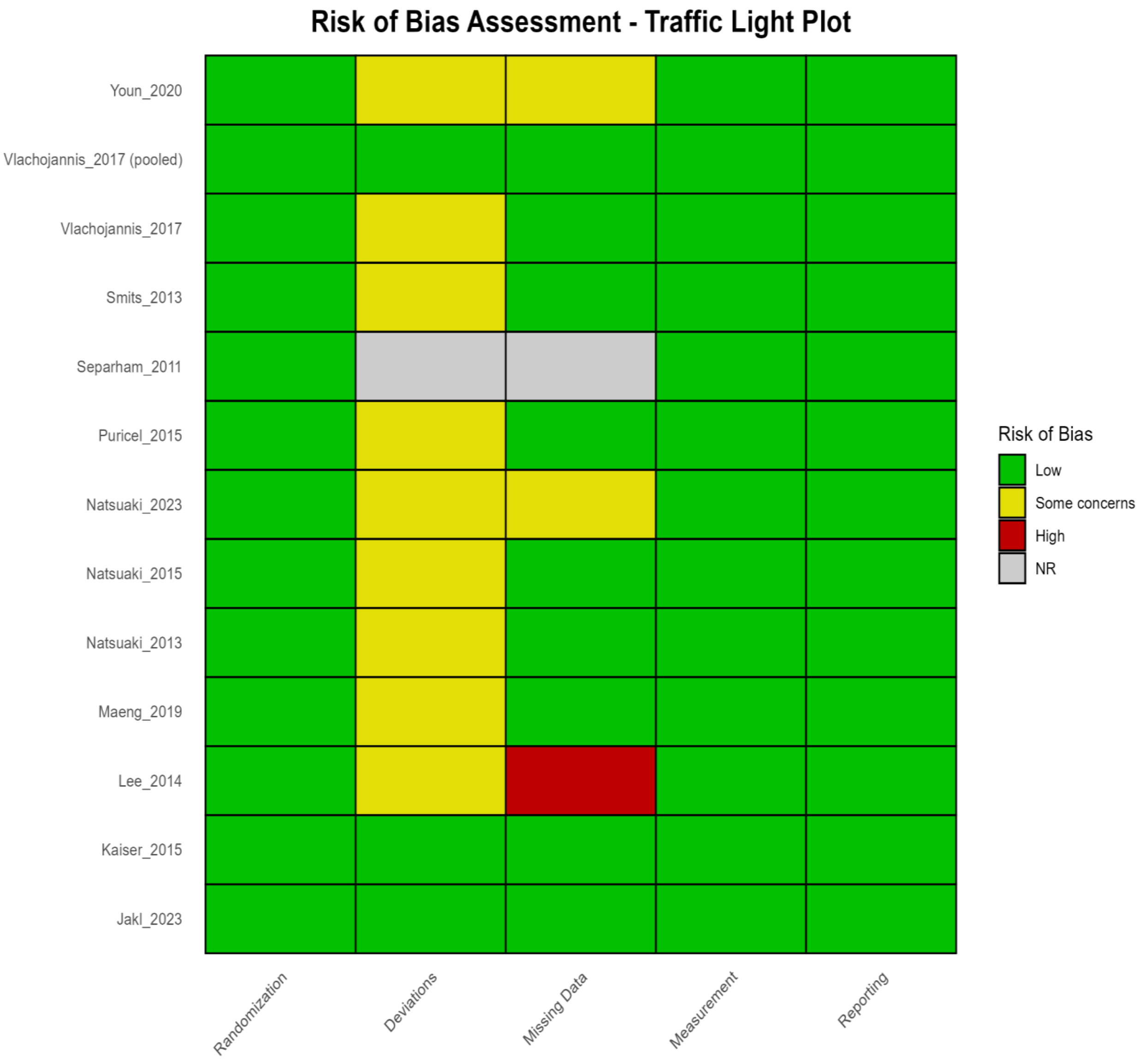


**Figure Legend:** Summary of risk of bias assessment using the Cochrane RoB 2 tool across five domains: (D1) randomization process, (D2) deviations from intended interventions, (D3) missing outcome data, (D4) measurement of the outcome, and (D5) selection of the reported result. Green indicates low risk of bias, yellow indicates some concerns, and red indicates high risk of bias.

**Supplementary Figure 2. Forest Plot of Device-Oriented Composite Endpoint**

| Study | EES Events | EES Total | BES Events | BES Total | RR | 95% CI | Weight |
|---|---|---|---|---|---|---|---|
| Natsuaki 2015 | 187 | 1618 | 196 | 1617 | 0.95 | [0.79; 1.15] | 14.8% |
| Puricel 2015 | 11 | 80 | 4 | 80 | 2.75 | [0.91; 8.27] | 0.4% |
| Maeng 2019 | 55 | 1385 | 60 | 1379 | 0.91 | [0.64; 1.31] | 4.1% |
| Lee 2014 | 42 | 255 | 40 | 245 | 1.01 | [0.68; 1.50] | 3.3% |
| Vlachojannis 2017 (Pooled) | 301 | 2530 | 430 | 3412 | 0.94 | [0.82; 1.08] | 27.4% |
| Youn 2020 | 23 | 638 | 14 | 634 | 1.63 | [0.85; 3.14] | 1.2% |
| Natsuaki 2013 | 125 | 1618 | 129 | 1617 | 0.97 | [0.76; 1.23] | 9.4% |
| Natsuaki 2023 | 269 | 1213 | 290 | 1204 | 0.92 | [0.80; 1.06] | 24.7% |
| Smits 2013 | 34 | 912 | 76 | 1795 | 0.88 | [0.59; 1.31] | 3.3% |
| Vlachojannis 2017 (5yr) | 105 | 912 | 240 | 1795 | 0.86 | [0.69; 1.07] | 11.3% |
| **Random effects model** | | **11161** | | **13778** | **0.94** | **[0.88; 1.01]** | **100.0%** |

Heterogeneity: $I^2 = 0.0\%$, $\tau^2 = 0$, $\chi^2_9 = 7.43$ ($p = 0.5921$)

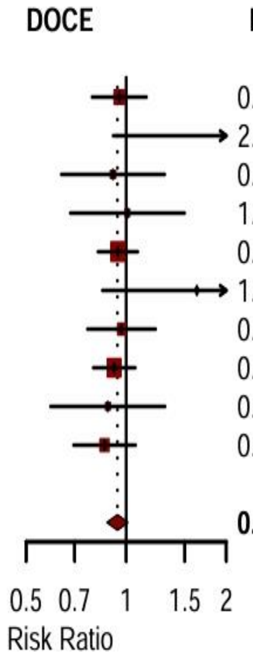


**Figure Legend:** Random-effects meta-analysis of the device-oriented composite endpoint (cardiac death, target vessel myocardial infarction, and target lesion revascularization) comparing everolimus-eluting stents with biolimus-eluting stents.

**Supplementary Figure 3. Leave-One-Out Sensitivity Analysis**

| Study | RR | 95%-CI | P-value | Tau2 | Tau | I2 |
|---|---|---|---|---|---|---|
| Omitting Jakl 2023 (9mo) | 0.93 | [0.87; 1.00] | 0.0497 | 0 | 0 | 0% |
| Omitting Jakl 2023 (60mo) | 0.93 | [0.87; 1.00] | 0.0588 | 0 | 0 | 0% |
| Omitting Natsuaki 2015 | 0.92 | [0.85; 0.99] | 0.0267 | 0 | 0 | 0% |
| Omitting Puricel 2015 | 0.92 | [0.86; 0.99] | 0.0273 | 0 | 0 | 0% |
| Omitting Maeng 2019 | 0.94 | [0.87; 1.02] | 0.1275 | 0 | 0 | 0% |
| Omitting Lee 2014 | 0.93 | [0.87; 1.00] | 0.0503 | 0 | 0 | 0% |
| Omitting Vlachojannis 2017 (Pooled) | 0.95 | [0.87; 1.02] | 0.1531 | 0 | 0 | 0% |
| Omitting Youn 2020 | 0.93 | [0.86; 0.99] | 0.0350 | 0 | 0 | 0% |
| Omitting Kaiser 2015 | 0.93 | [0.87; 1.00] | 0.0631 | 0 | 0 | 0% |
| Omitting Natsuaki 2013 | 0.93 | [0.86; 1.00] | 0.0604 | 0 | 0 | 0% |
| Omitting Natsuaki 2023 | 0.93 | [0.86; 1.01] | 0.0935 | 0 | 0 | 0% |
| Omitting Separham 2011 | 0.93 | [0.87; 1.00] | 0.0555 | 0 | 0 | 0% |
| Omitting Smits 2013 | 0.93 | [0.87; 1.00] | 0.0633 | 0 | 0 | 0% |
| Omitting Vlachojannis 2017 (5yr) | 0.94 | [0.87; 1.01] | 0.1027 | 0 | 0 | 0% |
| **Random effects model** | **0.93** | **[0.87; 1.00]** | **0.0525** | **0** | **0** | **0%** |

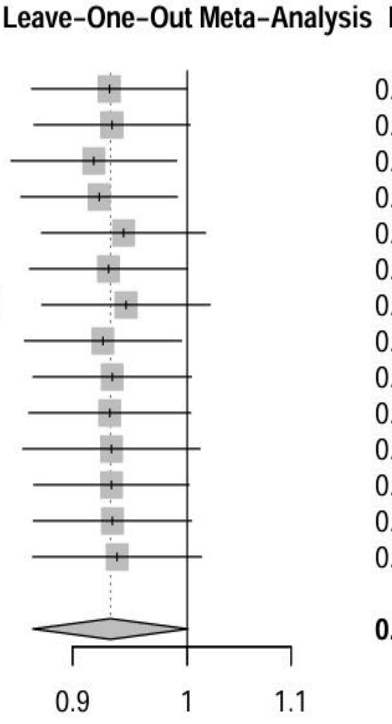


**Figure Legend:** Sequential exclusion of each study from the meta-analysis of major adverse cardiac events, showing the recalculated pooled risk ratio and 95% confidence interval. The consistency of results demonstrates the robustness of the findings.

**Supplementary Figure 4. Cumulative Meta-Analysis**

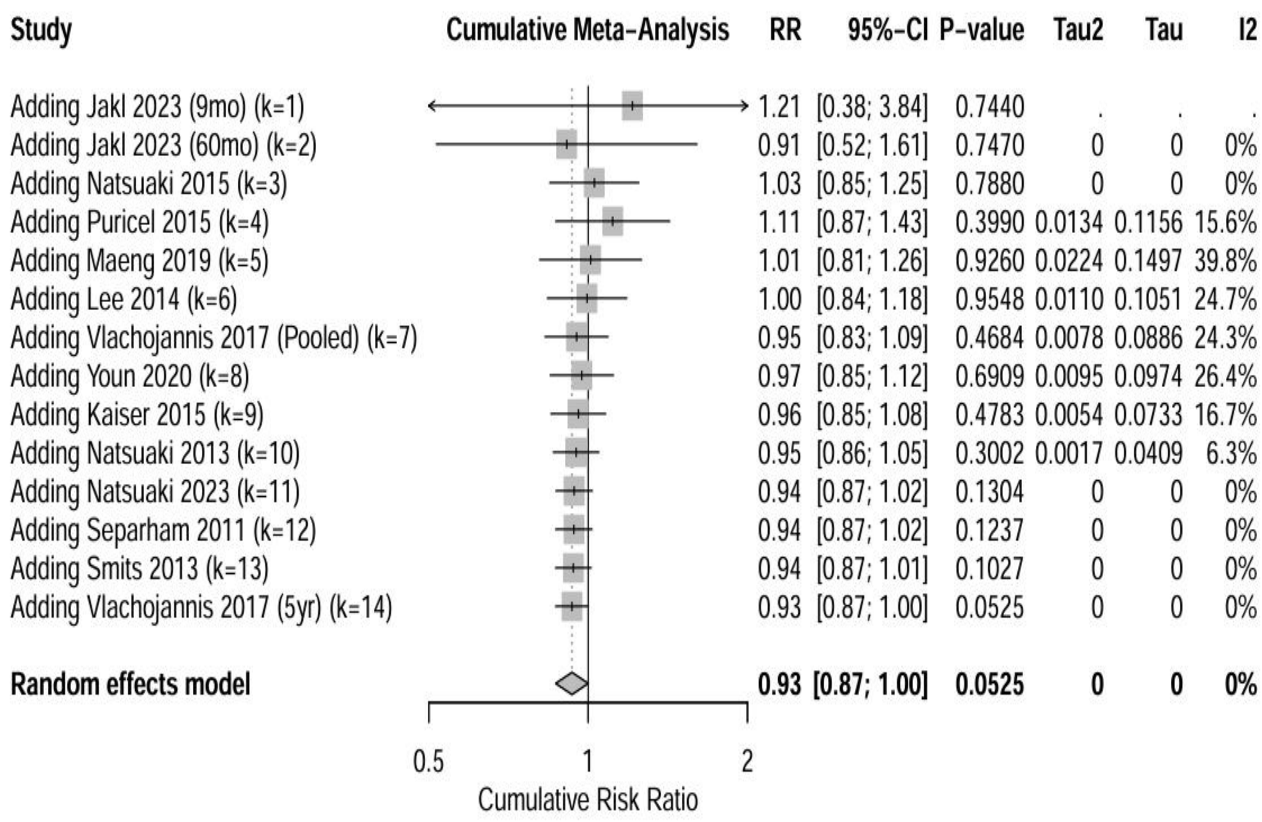


**Figure Legend:** Cumulative meta-analysis of major adverse cardiac events ordered by publication year, showing the evolution of the pooled effect estimate as studies were sequentially added. The effect estimate stabilized after approximately 10,000 patients were included.
